# Highly adaptive LASSO: Machine learning that provides valid nonparametric inference in realistic models

**DOI:** 10.1101/2024.10.18.24315778

**Authors:** Zachary Butzin-Dozier, Sky Qiu, Alan E. Hubbard, Junming (Seraphina) Shi, Mark J. van der Laan

**Affiliations:** Department of Biostatistics University of California, Berkeley Berkeley, CA 94704

**Keywords:** Machine Learning, Causal Inference, Targeted Learning

## Abstract

Understanding treatment effects on health-related outcomes using real-world data requires defining a causal parameter and imposing relevant identification assumptions to translate it into a statistical estimand. Semiparametric methods, like the targeted maximum likelihood estimator (TMLE), have been developed to construct asymptotically linear estimators of these parameters. To further establish the asymptotic efficiency of these estimators, two conditions must be met: 1) the relevant components of the data likelihood must fall within a Donsker class, and 2) the estimates of nuisance parameters must converge to their true values at a rate faster than *n*^−1/4^. The Highly Adaptive LASSO (HAL) satisfies these criteria by acting as an empirical risk minimizer within a class of *càdlàg* functions with a bounded sectional variation norm, which is known to be Donsker. HAL achieves the desired rate of convergence, thereby guaranteeing the estimators’ asymptotic efficiency. The function class over which HAL minimizes its risk is flexible enough to capture realistic functions while maintaining the conditions for establishing efficiency. Additionally, HAL enables robust inference for non-pathwise differentiable parameters, such as the conditional average treatment effect (CATE) and causal dose-response curve, which are important in precision health. While these parameters are often considered in machine learning literature, these applications typically lack proper statistical inference. HAL addresses this gap by providing reliable statistical uncertainty quantification that is essential for informed decision-making in health research.

## 1 Introduction

Understanding the treatment effect on health-related outcomes requires one to first define a causal parameter of interest. The causal parameter is often a feature(s) of the probability distributions in the unseen counterfactual scenario. To estimate the causal parameter using observed data, one needs identification assumptions to turn the causal parameter into a statistical estimand (Pearl [2000], Pearl et al. [2016]). The remaining work is purely statistical, as the investigator constructs estimators for those estimands. Most of the semiparametric methodological research has focused on constructing asymptotically linear estimators of smooth (formally, pathwise differentiable) parameters of the data distribution. These are estimators that can be represented, up to a negligible term, as independent and identically distributed (i.i.d) random variables with mean zero, called influence curves (Bickel et al. [1993]). Such methods include one-step estimation (Bickel et al. [1993]), estimation equation approaches (Robins et al. [1995], van der Laan and Robins [2003], Chernozhukov et al. [2018]) as well as plug-in estimators like the targeted maximum likelihood estimator (TMLE) (van der Laan and Rubin [2006], van der Laan and Rose [2011, 2017]). Due to its plug-in property, TMLE respects any global constraints on the parameter of interest. For example, if one is interested in estimating the difference in survival probability under an active treatment versus the standard-of-care, then the estimate produced by TMLE is guaranteed to fall between −1 to 1. Other plug-in estimators (such as *g*-computation) typically are not asymptotically linear in nonparametric/semiparametric statistical models (Robins [1986]). These estimators all rely on the estimation of relevant nuisance parameters of the data-generating distribution (DGD). Take the TMLE for the average treatment effect (ATE) as an example, the two nuisance parameters are the conditional mean of the outcome given treatment and covariates (outcome regression) and the treatment probability given covariates (propensity score). Theoretical analysis of these estimators suggests that for them to be asymptotically linear and efficient, the nuisance parameters generally need to be estimated at a rate faster than *n*^−1/4^ in *L*^2^(*P*_0_)-norm, with *n* being the sample size (van der Laan [2015]). Under parametric models, one is often able to attain such rate. However, one risks introducing model misspecification bias. Under knowledge and empirical evidence-based realistic models (which are often nonparametric or semiparametric), the rate is challenging to attain due to *curse-of-dimensionality*. Consequently, one needs flexible machine learning algorithms that are not only guaranteed to be consistent but also be able to approximate the true target function at the required rate in the observed sample size. It is therefore an open problem in semiparametric efficiency theory of how to thread the needle of having machine learning algorithms that are flexible enough to accurately estimate relevant factors of the data likelihood, yet not so volatile that they violate the conditions required to establish asymptotic linearity and efficiency (Robins and Ritov [1997]).

In recent years, the Highly Adaptive LASSO (van der Laan [2015], Benkeser and van der Laan [2016]) was proposed as a machine learning algorithm that is flexible enough to capture any realistic function one would encounter in health-related fields *and* satisfies the theoretical conditions of asymptotically linear and efficient estimators. HAL is an ideal machine learning algorithm in causal inference tasks for the following three reasons. First, it has been empirically demonstrated to be competitive in predictive accuracy against alternative algorithms including regression trees, random forests, and gradient-boosted machines (Benkeser and van der Laan [2016]). Second, it has been theoretically proven to satisfy the convergence rate condition for constructing asymptotically linear and efficient estimators (Bibaut and van der Laan [2019]). Third, the model fit of HAL is a linear combination of basis functions that resembles fits obtained using parametric modeling approaches and is easily interpretable, making it conducive to decision-making in public health and medicine. Beyond having ideal properties of machine learning algorithms, the working model implied by a HAL fit also provides a foundation for robust inference for non-pathwise differentiable parameters, such as the conditional average treatment effect (CATE) and dose-response curve for a continuous treatment, which are of significant interest in areas of precision health (van der Laan [2023], Shi et al. [2024]). Although these parameters are discussed in machine learning and deep learning literature (Künzel et al. [2019], Nie and Wager [2021], Curth and van der Schaar [2021]), their applications generally lack valid statistical inference. However, in health research, statistical inferences (reliable measures of estimation uncertainty) are central to making informed, strategic decisions about new strategies of care.

This paper is organized as follows. In Section 2, we introduce the fundamental concepts necessary to understand and construct asymptotically linear estimators, along with the conditions required to establish their asymptotic efficiency. In Section 3, we explore a specific class of functions, known as *càdlàg* functions, and their representations, which provide the foundation for the motivation and development of HAL. Section 4 covers practical applications of HAL, detailing various ways it can be utilized in causal inference problems for estimation and robust statistical inference, including in more challenging scenarios involving non-pathwise differentiable parameters. We conclude with a summary and discussion on future research directions, including the computational aspects of HAL and the exciting opportunities it presents. We have included a GitHub repository that provides R code for a potential application of these methods (https://raw.githack.com/SeraphinaShi/HAL-in-R/main/Demo.html).

## 2 Preliminaries

In this section, we introduce the preliminaries of HAL. Namely, we start by considering a simple point-treatment causal inference scenario where one is interested in estimating the average treatment effect. Using this example, we introduce the basics of asymptotically linear estimators. Then, we discuss an important theoretical framework, efficiency theory, that supports the construction and analysis of efficient estimators. Discussions in this section are not comprehensive, but rather aim to foreshadow the key theoretical properties of HAL that will be explored in subsequent sections.

Let *Pf* =∫ *f* (*o*)*dP* (*o*) and 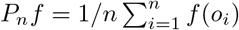 for a function *f* (*o*). Consider the simplest point-treatment causal inference setting where our observed data consists of *n* copies of the random variable *O* = (*W, A, Y*) that are drawn i.i.d from the true data-generating distribution *P*_0_ ∈ℳ, where ℳ is the statistical model. In particular, *W* ∈ ℝ^*d*^ is a vector of baseline covariates, *A*∈ {0, 1*}* is a binary treatment, and *Y* ∈ ℝ is the outcome of interest. Suppose we have a one-dimensional target parameter Ψ(*P*) ∈ ℝ of interest, which can be viewed as a mapping from any probability distribution in the statistical model to a real-value. Consider the average treatment effect (ATE) parameter defined as Ψ(*P*) = *E*(*Y*_1_ − *Y*_0_), where *Y*_1_ and *Y*_0_ are the potential outcomes under the hypothetical treatment and control, respectively. We will use the ATE parameter as a running example in this paper. We assume a structural causal model specified by *W* = *f*_*W*_ (*U*_*W*_), *A* = *f*_*A*_(*W, U*_*A*_), and *Y* = *f*_*Y*_ (*W, A, U*_*Y*_), where *U* = (*U*_*W*_, *U*_*A*_, *U*_*Y*_) are exogenous variables, and we left the functional forms of *f*_*W*_, *f*_*A*_, and *f*_*Y*_ unspecified. Counterfactual distributions are defined by deterministically setting the treatment node *A* to 0 or 1. Additionally, we assume no unmeasured confounding or violations of positivity, which are standard identification assumptions in causal inference that allow the causal parameter to be expressed as a statistical estimand given by Ψ(*P*_0_) = *E*_0_[*E*_0_(*Y* |*W, A* = 1) −*E*_0_(*Y* |*W, A* = 0)] (Tian and Pearl [2002], Hernán and Robins [2010], Dang et al. [2023]).

### 2.1 mAsymptotically linear estimators

An estimator 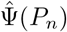 of the true ATE *ψ*_0_ ∈ ℝ is asymptotically linear with influence curve *D*(*P*_0_)(*O*) if it can be represented as the empirical mean of the influence curve plus a term that converges in probability to zero at a rate faster than 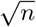 (Bickel et al. [1993]). The asymptotic variance of such an estimator is determined by the variance of the influence curve. This naturally raises the question of whether there exists an optimal influence curve that minimizes the asymptotic variance. The influence curve with the smallest asymptotic variance is referred to as the efficient influence curve, and estimators that are asymptotically linear with this influence curve are called asymptotically linear and efficient among a class of regular estimators. These estimators are ideal for inference because they achieve the smallest asymptotic variance, leading to the least uncertainty. An asymptotically linear and efficient estimator takes the form:

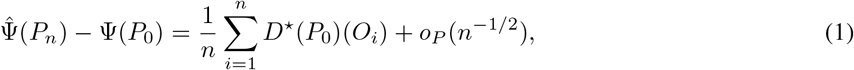

where *D*^⋆^(*P*_0_)(*O*) is the efficient influence curve.

The recipe to construct such an estimator for the parameter of interest typically involves identifying the canonical gradient of the target parameter, which is a mathematical object that can be derived at distribution *P* ∈ *ℳ*. The canonical gradient corresponds to the efficient influence curve in the context of asymptotically linear estimators. In our example, the canonical gradient of the ATE parameter Ψ at *P* in a nonparametric statistical model is given by

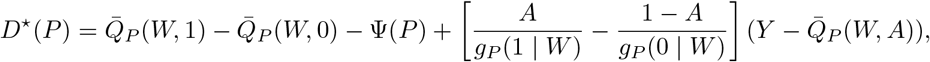

where 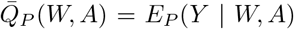 (a.k.a outcome regression) and *g*_*P*_ (*a* | *W*) = *P* (*A* = *a* | *W*) (a.k.a treatment mechanism) for *a* ∈ *{*0, 1*}*.

### 2.2 Efficiency theory

Suppose 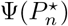 is an estimator of the true ATE estimand Ψ(*P*_0_) constructed by TMLE, it then has the property that 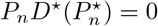. In other words, TMLE (and some other debiasing machine learning techniques in causal inference) attempts to solve the empirical mean of the efficient influence curve. Now, to see the conditions under which Equation (1) holds for 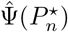, we can analyze the difference between the TMLE estimator and the true value of the estimand. Define the exact remainder as

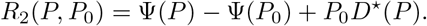

When Ψ is pathwise differentiable, we have

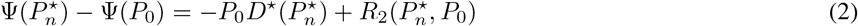

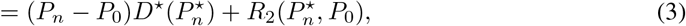

where the second line follows from 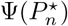 being a TMLE so 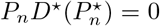. The first term on the right-hand side of Equation (3) can be controlled by applying empirical process theory. In particular, assume that 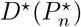 falls in a *P*_0_-Donsker class with probability tending to 1, and that 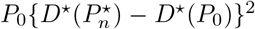 converges in probability to zero. Then,

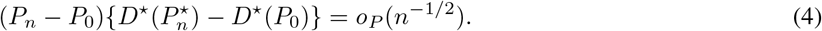

This result is also known as the asymptotic equicontinuity (van der Vaart and Wellner [1996]). Therefore, with (4), we can rewrite Equation (3) as

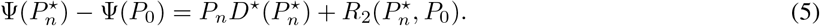

This is one step closer to our desired form of asymptotically linearity and efficiency in Equation (1). We also need to make sure that the exact remainder term diminishes fast enough. Note that, one could analytically derive the closed-form of the exact remainder term *R*_2_(*P, P*_0_) by its definition. In the case of ATE, the remainder is

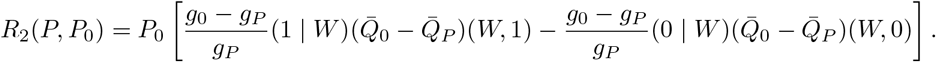

This implies that if the estimated nuisance parameters converge to their truths at a rate faster than *n*^−1/4^ in *L*^2^(*P*_0_)-norm, then by the Cauchy–Schwarz inequality we have 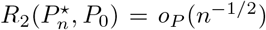. Together with the Donsker class and empirical process condition, we have

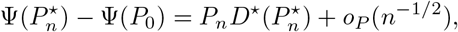

thereby establishing the asymptotic linearity and efficiency of the estimator 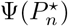.

To summarize, for a TMLE estimator to achieve efficiency, two conditions must be met: 1) 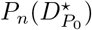to fall in a Donsker class (or the relevant components of *P*_0_ fall in a Donsker class), and 2) 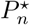 to converge to *P*_0_ at a rate such that 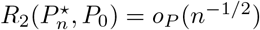. In our example, these requirements translate to the need for both the outcome regression and the treatment mechanism to fall within a Donsker class and for their estimates to converge to the true values at a rate of at least *n*^−1/4^ with respect to the square-root of the loss-based dissimilarity.

The first condition, concerning the Donsker class, can be bypassed using cross-fitting. Originally proposed for the classic one-step estimator, cross-fitting has since been incorporated into and combined with other approaches, including estimating equations and CV-TMLE (Hubbard and van der Laan [2009], Zheng and van der Laan [2010]). Cross-fitting allows us to avoid the Donsker class constraints and has been shown in realistic simulation studies to be a robust method with generally good performance, regardless of the adaptiveness of the estimators of the data-generating distribution (DGD) (Li et al. [2022]). However, this approach comes with a trade-off, as it expands the model to one without Donsker constraints, making it more challenging to control the exact remainder term at the desired rate. Intuitively, the goal is to identify a function class that is neither too large nor too restrictive, yet flexible enough to include functions encountered in practice. *Càdlàg* functions with bounded sectional variation norms meet these criteria and also happen to form a Donsker class Zheng and van der Laan [2010].

## 3 Highly Adaptive LASSO

In this section, we start by introducing a particular function class that has properties satisfying the conditions for asymptotic linearity and efficiency. In particular, we discuss a general representation of functions in this class that motivate HAL. To close this section, we share some implementation details of HAL.

### 3.1 *Càdlàg* function and its representation

Highly Adaptive LASSO is an empirical risk minimizer in a function class consisting of *càdlàg* functions with bounded sectional variation norm *M*. We use ℱ_*M*_ ([0, *τ* ]^*d*^) for *τ >* 0 to represent this function class. *Càdlàg* (*continue à droite, limite à gauche*) is an acronym for a phrase in French that is translated as right-continuous with left limits. In this subsection, we introduce key characteristics of functions in this class that motivate the use of HAL. We define the sectional variation norm, ||*f*||_*v*_, of a multivariate function *f* as

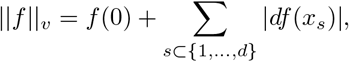

where *f* (*x*_*s*_) denotes the function *f* where the coordinates not in the set *s* are set to 0. This norm measures the total variation of a function, indicating how much the function fluctuates across all coordinates of its domain. Any *càdlàg* function *f* with a bounded sectional variation norm ||*f*||_*v*_ can be represented as follows (Gill et al. [1995]):

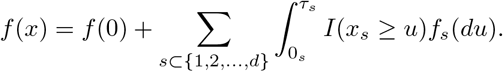

This representation can be approximated by:

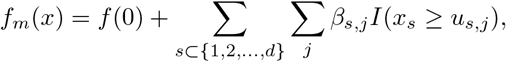

where *{ u*_*s,j*_ : *s, j }* is a finite set of knot points that sets coordinates not in *s* to zeros (Benkeser and van der Laan [2016]). *I*(*Z*) is the indicator variable that equals to 1 if statement *Z* is true. Rather than minimizing the empirical risk over all *càdlàg* functions, HAL focuses on functions with discrete support points, which form a sieve within the parameter space. This sieve is large enough to ensure that the approximation error of the true function *f* is minimal, allowing us to focus on estimating the minimizer of the true risk within this finite-dimensional model. Such sieves also enhance finite sample performance.

There are various ways to choose the knot points, and HAL specifically uses a set of data-dependent knot points that are uninformative, meaning they are determined by the observed data. We will discuss constructing this set of knot points in the subsequent subsection. To summarize, any *càdlàg* function can be approximated as a linear combination of indicator basis functions, a specification known as 0th-order HAL. The sectional variation norm of the function is equivalent to the *L*_1_-norm of all the coefficients *β*. Therefore, 0th-order HAL solves the following constrained optimization problem:

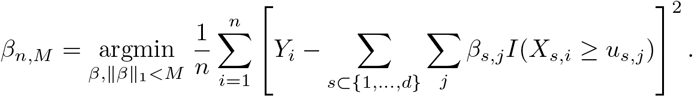

The loss function considered here is the squared-error loss, which is well-suited for continuous outcomes. However, HAL can be paired with any scientifically relevant loss function depending on the context. For instance, when the outcome is binary, the log-likelihood loss—equivalent to using the logistic link in generalized linear models—is appropriate. Another example is when estimating conditional densities. In this case, it is recommended to parameterize the conditional density using conditional hazard functions, as detailed in Rytgaard et al. [2023] and Munch et al. [2024].

Now, if we use HAL to estimate the nuisance parameters, e.g. 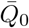 and *g*_0_, what assumptions do we impose on their true forms? The first assumption states that 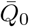 and *g*_0_ are *càdlàg* functions. The second assumption is that the variation norm of 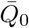 and *g*_0_ is finite. These assumptions impose global constraints on the true function, rather than the local smoothness constraints often encountered in current literature (such as requiring a certain number of derivatives) that are typically necessary to ensure sufficient rates of convergence. Additionally, the assumptions we made allow the function to have jumps. Such a constraint is rarely violated in health data settings, as the mechanisms generating health data across various data types and study designs generally have inherent limitations on the volatility of the resulting functions, which almost always have finite bounds, both in terms of values and changes. The class of *càdlàg* functions also happens to be a Donsker class with a bounded entropy integral. Therefore, it satisfies the Donsker class condition in establishing the asymptotic efficiency discussed in Section 2. The fact that *càdlàg* functions belong to the Donsker class is also the reason one could successfully establish their fast rate of convergence.

### 3.2 Implementation of Highly Adaptive LASSO

This constrained optimization problem is equivalent to LASSO, where one seeks to minimize the penalized *L*_1_-norm loss function (Tibshirani [1996]). The only difference is that instead of inputting the original design matrix *X* into LASSO, we input a HAL design matrix with indicator basis functions and run the same optimization algorithm. The set of knot points used by HAL is implied by the data. Specifically, for each subset *s* of the covariate index set {1, 2, …, *d}*, we construct up to *n* basis functions, where the *i*-th basis function is *I*(*X*_*i*_ ≥ *x*_*s,i*_), with *x*_*s,i*_ denoting the *i*-th realization of *X* and variables with indices in *s* set to zero. These basis functions are conveniently formed by 1) tabulating knot points for each variable at every observed data point, i.e., *I*(*X*_1_ ≥ *x*_1,1_), …, *I*(*X*_1_ ≥ *x*_1,*n*_), …, *I*(*X*_*d*_ ≥ *x*_*d*,1_), …, *I*(*X*_*d*_ ≥ *x*_*d,n*_), and 2) taking all two-way, three-way, and up to *d*-way tensor products of these one-way indicator basis functions. The resulting basis set can thus be seen as an exhaustive collection of indicator basis functions, including both main effects and interaction terms. The R package *hal9001* implements this procedure by providing functions to enumerate and evaluate the basis functions, and then calling the *glmnet* R package to solve the LASSO Hejazi et al. [2020], Hastie et al. [2021]. Figure 1 illustrates the use of HAL with 0th-order spline basis functions to fit a univariate sinusoidal function. HAL is flexible enough to fit the true function well, while the cross-validation-selected total variation norm effectively prevents overfitting. Figure 1 shows three fits with variation norms of 4.8, 13.4, and 35.2. The optimal fit, with a variation norm of 13.4, is chosen using cross-validation and is close to the true *L*_1_-norm of 16.

**Figure 1.**
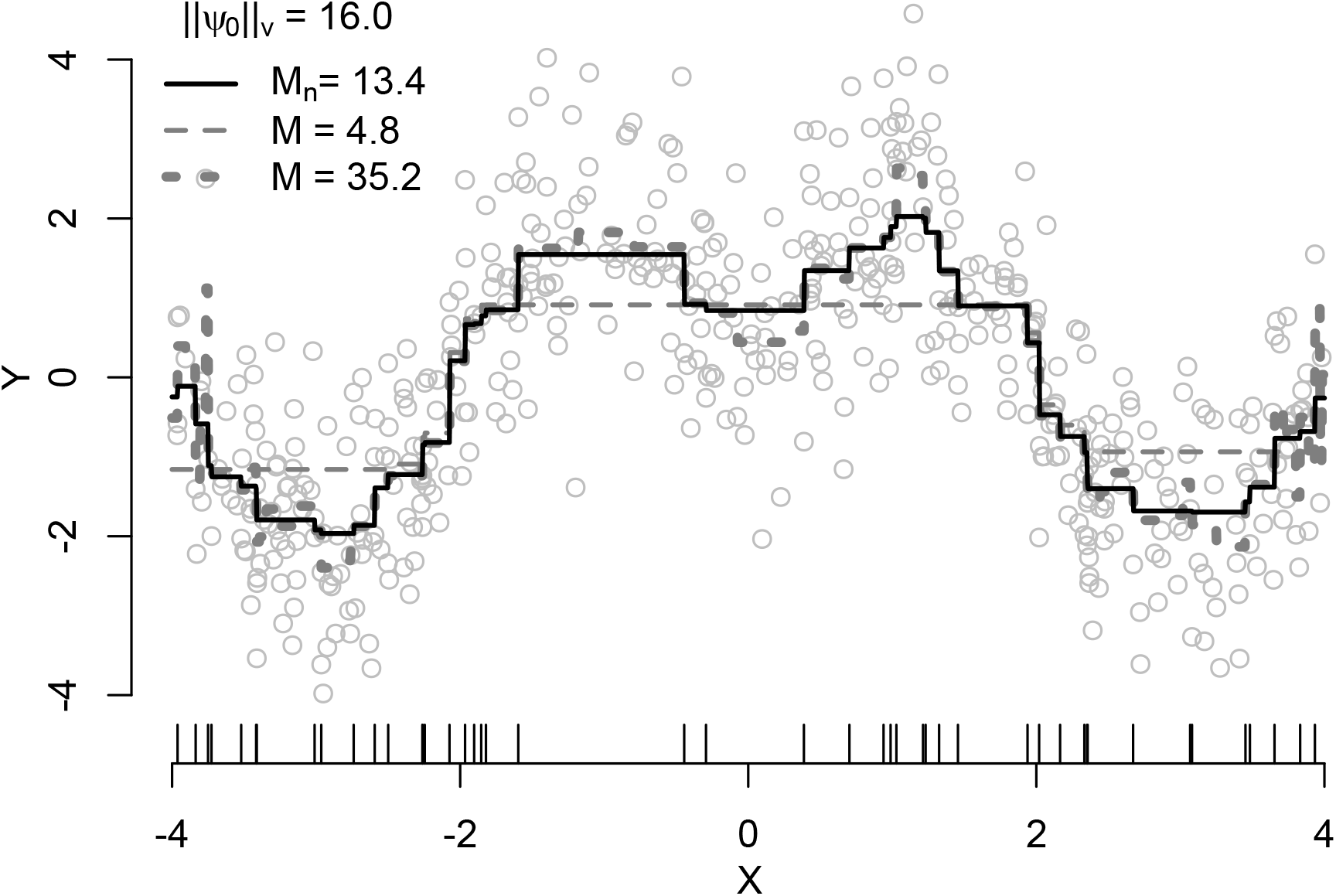
HAL in the univariate setting. Benkeser and van der Laan [2016] conducted a simulation, drawing 500 independent copies of *X* from a Uniform(− 4, 4) distribution and 500 copies of *ϵ* from a Normal(0,1) distribution where *Y* = 2*sin*(*π/*2 |*X*|) + *ϵ* so that ||*ψ*_0_|| _*υ*_. The HAL basis functions included *n* indicators at observed data points 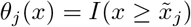 for *j* = 1, …, *n* and selected the bound on the variation norm via ten-fold cross-validation from 100 possible bounds from 0 to 350.

## 4 Highly Adaptive LASSO in practice

In this section, we begin by considering the use of HAL within the TMLE framework, which ensures the efficiency of estimators. We then discuss the application of nonparametric bootstrap methods for HAL-TMLE, which improve the finite sample coverage of confidence intervals. The fact that HAL-TMLE solves many score equations also motivates the development of HAL-based plug-in estimators, which, when undersmoothing is applied, can achieve efficiency independently of the TMLE framework. Additionally, we summarize the pointwise asymptotic normality results of higher-order HAL, enabling its use in estimating non-pathwise differentiable parameters and obtaining valid statistical inference within its data-adaptively learned working model. Finally, we cover recent advancements in HAL within the adaptive-TMLE framework, noting its advantages in scenarios with empirical positivity violations and its applications in data augmentation, such as integrating randomized controlled trial data with real-world observational data.

### 4.1 HAL-based TMLE

Previous work has demonstrated HAL’s competitive performance in machine learning tasks compared to algorithms such as regression trees, random forests, and gradient-boosted machines (Benkeser and van der Laan [2016]). The benefits of HAL extend far beyond these tasks. HAL can be used within the general TMLE framework as a nonparametric regression method for estimating nuisance parameters. When HAL is used to estimate these parameters, the resulting TMLE estimator of a pathwise differentiable parameter will be efficient within a statistical model assuming the true nuisance functions are *càdlàg* with finite sectional variation norms (van der Laan [2017]). As discussed previously, this assumption is generally satisfied for most functions in health sciences. The fast convergence rate of HAL ensures that the exact remainder term (defined in Section 2.2) diminishes faster than *n*^−1/2^, which is crucial for establishing asymptotic efficiency. Specifically, HAL with higher order splines has even faster rates of *n*^−(*k*+1)*/*(2 *k* +3)^ up till a power of log *n* depending on the dimension, where *k* = 0, 1, … is the assumed order of smoothness (van der Laan [2023]). Moreover, IPTW estimators can also be made efficient when HAL is used to estimate the propensity score, particularly when undersmoothing is applied (Ertefaie et al. [2023]). Undersmoothing involves selecting an *L*_1_-norm larger than the one chosen by cross-validation, ensuring that the resulting IPTW estimator of the target parameter solves a specific efficient influence curve equation. The need for undersmoothing arises for two reasons. First, due to the penalization on the *L*_1_-norm of the coefficients in a HAL fit, it only solves score equations approximately, rather than exactly like a maximum likelihood estimator (MLE). Second, even with a relaxed-HAL (taking the basis functions with non-zero coefficients and fitting an MLE), although one solved a set of score equations corresponding with the basis functions and their linear span, one also needs undersmoothing to ensure that the linear span of the scores approximates the score in the EIC, e.g. (2*A* − 1)*/g*(*A* | *W*) in the case of ATE, at a rate *n*^−1/4^. From a practical perspective, HAL can be fine-tuned in several ways. If one has prior knowledge regarding the form of the true underlying function, they can incorporate this information in HAL’s hyperparameters. For example, if the true functional form includes a specific additive function, those basis functions can be added without having their coefficients considered in the *L*_1_-penalization. HAL can also specify scenarios where certain basis functions are known to have monotonicity constraints. If the true function is likely to exhibit higher-order smoothness, one can opt for higher-order HAL instead of 0th-order spline basis functions. Even in the absence of such prior knowledge, different specifications of HAL can still be used as candidates within a super learner, with cross-validation employed to select the best among them. The benefit of including several different specifications of HAL in a super learner library is that the super learner will perform as well as the best candidate learner among all HAL estimators (van der Laan et al. [2007]). Therefore, at a minimum, the super learner library will achieve the convergence rate of *n*^−1/3^(log *n*)^*d/*2^. Hence, if used as an estimator for nuisance parameter estimation in our running example of estimating the ATE, the convergence rate condition will be satisfied, and a TMLE estimator with HAL used to estimate nuisance parameters will be asymptotically linear and efficient.

### 4.2 Nonparametric bootstrap inference for HAL-TMLE

With a HAL-TMLE estimator, statistical inference can be made by leveraging its asymptotic normality. However, inference can still be challenging in finite sample settings. In certain settings, the higher-order remainder may easily dominate the first-order term (i.e. the empirical mean of the efficient influence curve), leading to an underestimation of the variance and low coverage probability of the resulting confidence intervals. To address this, the nonparametric bootstrap method enables valid statistical inferences in HAL-TMLE (Cai and van der Laan [2020]). Variance estimation based on the nonparametric bootstrap has been shown in simulations to improve confidence interval coverage compared to first-order Wald-type confidence intervals (see Figure 2).

**Figure 2.**
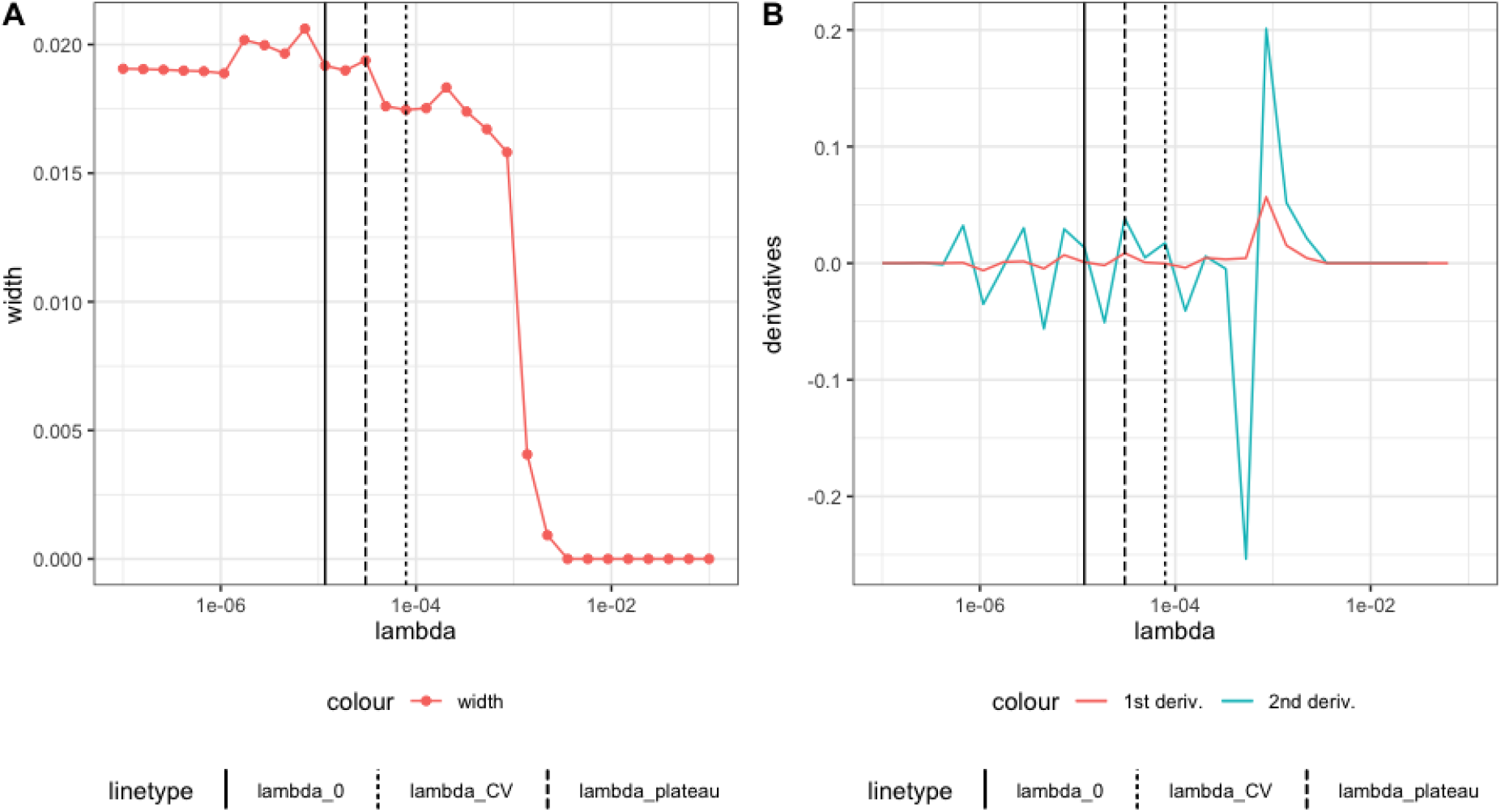
A. Simulation of Wald interval as a function of *λ*. B. First-order and second-order derivatives of the curve. The solid line represents *λ*_0_, the fine dotted line represents *λ*_*CV*_, and the thick dotted line represents *λ*_*plateau*_.(Reproduced figure Cai and van der Laan [2020]).

Constructing nonparametric bootstrap confidence intervals is fairly straightforward. For each nuisance parameter, one begins by fitting a HAL model, selecting the *L*_1_-norm penalty *λ* using *V* -fold cross-validation. A relaxed-HAL is then constructed by taking the HAL basis functions with non-zero coefficients (the LASSO-selected support set) and running an unpenalized regression. These initial estimates are used in the TMLE targeting step to obtain an updated estimate of the target parameter. For each HAL fit with *λ* in a pre-specified grid of *λ* values, where the *L*_1_-norm is greater than the CV-selected *L*_1_-norm, bootstrap sampling is performed to obtain nonparametric bootstrap confidence intervals for the TMLE estimate. The widths of these intervals are plotted, and the interval corresponding to the *λ* value at which the widths start to plateau is selected as the final bootstrap confidence interval. This plateauing behavior indicates that the interval widths increase monotonically and stabilize when the fit starts approximating the true regression function nicely, where they approach a plateau. Figure 3 illustrates this in a simulation. Theoretical support for this behavior can be found in Theorem 1 of Davies and van der Laan [2014]. In practice, the plateau selector is implemented by finding the maximum of the second-order derivative of the confidence interval width.

**Figure 3.**
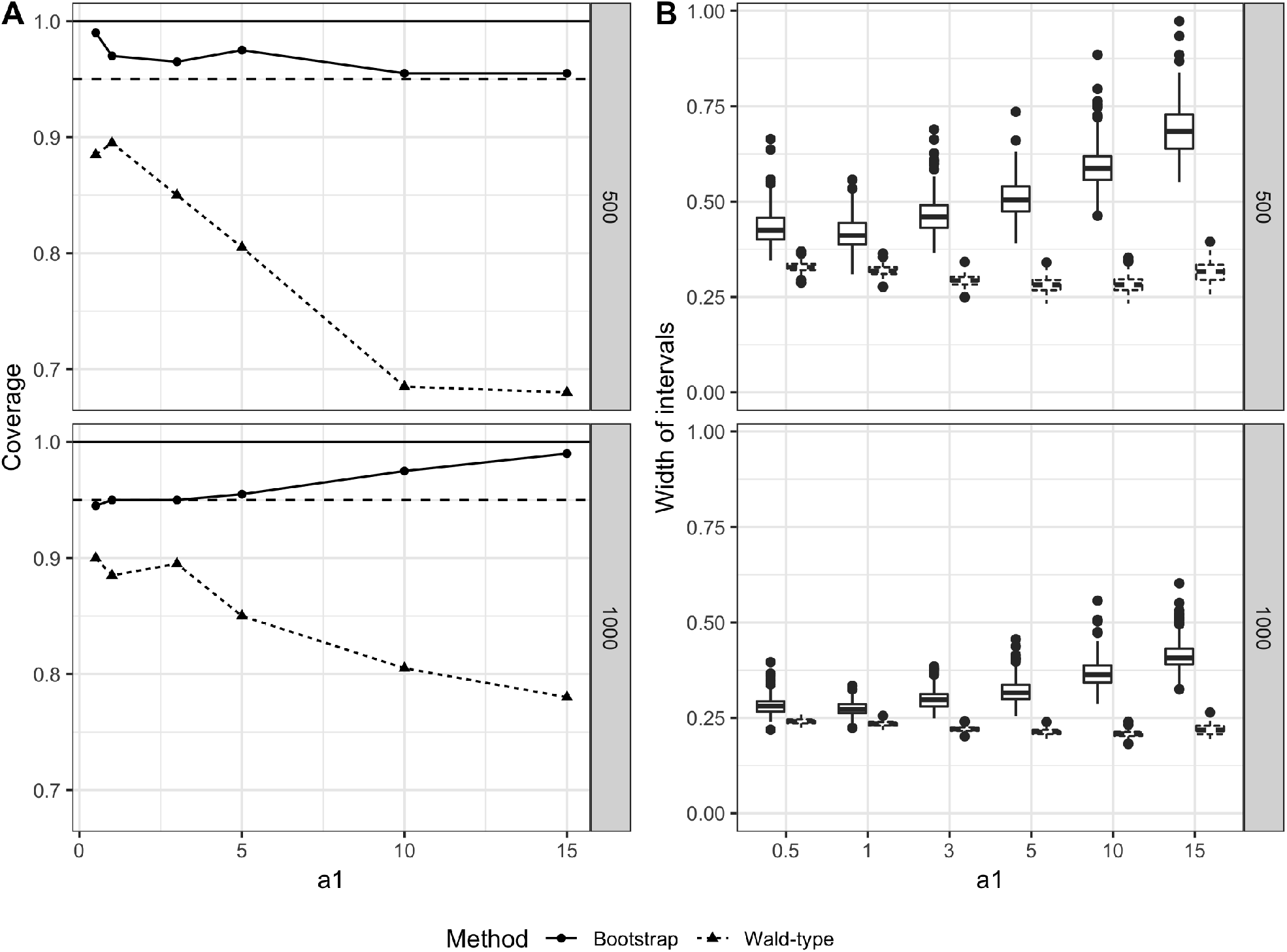
Average treatment effect using nonparametric bootstrap and Wald method for sectional variation norm of 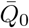 (Cai and van der Laan [2020]).

### 4.3 HAL-based plug-in estimator

Given the advantages of using HAL for estimating components of the data likelihood, one naturally wonders whether a plug-in estimator can be constructed directly from HAL for any pathwise differentiable parameter. For instance, in our ongoing example, if HAL is used to estimate the outcome regression, what are the properties of an estimator that simply takes the difference between HAL predictions evaluated at *A* = 1 and *A* = 0 and then averages the results? Interestingly, HAL can yield an asymptotically efficient estimator of any pathwise differentiable parameter when undersmoothing is applied (van der Laan et al. [2023a]). Undersmoothing involves “overfitting” the data in a beneficial way, leveraging the fact that MLEs solve a wide range of score equations. This process ensures that the score equations solved by the HAL-MLE closely approximate the score equation implied by the canonical gradient of the target parameter at the data distribution. In practice, undersmoothing is achieved by enlarging the *L*_1_-norm (equivalently, reducing the LASSO penalty parameter *λ*) such that the empirical mean of the efficient influence curve of the target parameter is solved at the level *σ*_*n*_*/*(*n*^1/2^ log *n*), where 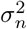 is the variance estimate based on the efficient influence curve. This approach is beneficial because an efficient estimator can be obtained directly from the HAL fit without needing an additional targeting step. If a targeting step is performed on the undersmoothed fit, the resulting estimate is unlikely to change considerably.

### 4.4 HAL for non-pathwise differentiable parameters

Beyond its use in TMLE and other doubly-robust estimation frameworks, HAL (with higher-order splines) has been shown to possess the property of pointwise asymptotic normality (van der Laan [2023]). This finding implies that HAL can provide valid statistical inference even for non-pathwise differentiable parameters, such as the causal dose-response curve. Although the asymptotic results in van der Laan [2023] were derived under the assumption that the smoothness order of the true function is known, in practice, a discrete super learner can select among candidate smoothness classes without sacrificing convergence rates. Specifically, due to the asymptotic equivalence of the *L*_1_-norm-specific cross-validation selector with the oracle selector, this discrete super learner can achieve a convergence rate as if the true smoothness had been known a priori (van der Laan [2023]).

Unlike the main-term parametric working models often used for convenience, the working model implied by HAL is learned data-adaptively and provides strong approximations of the true function being estimated. This data-adaptive approach allows HAL to offer a natural and straightforward way to conduct robust statistical inference using the classic delta method in parametric models. In many cases where statisticians previously relied on potentially misspecified parametric models for tractable inference, HAL-based data-adaptive working models now offer a powerful alternative. For example, due to its pointwise normality property, HAL can be used to construct simultaneous confidence bands for non-pathwise differentiable parameters like the causal dose-response curve (Shi et al. [2024]).

### 4.5 HAL for adaptive-TMLE

In recent work, HAL has been utilized as a key for working model selection under the adaptive-TMLE (A-TMLE) framework (van der Laan et al. [2023b]). A-TMLE is a recently proposed statistical estimation framework that regularizes an efficient TMLE of the ATE by adaptively performing a bias-variance trade-off. In scenarios where there are empirical positivity violations, estimating nonparametrically-defined target parameters can be challenging and may result in large variance estimates due to the inverse weighting of the treatment mechanism in the efficient influence curve. Rather than targeting the nonparametric parameter directly, A-TMLE focuses on a projection parameter, defined as the projection of the true data-generating distribution *P*_0_ onto a data-adaptively learned working model plugged in the original parameter mapping. While, in the limit, the working model can become fully nonparametric, in finite samples, A-TMLE adapts to the complexity of the working model and often provides more stable variance estimates. The approximation error of the projection estimand with respect to the true estimand is of second order, meaning that pursuing the projection parameter incurs no penalty for conducting valid statistical inference. In scenarios with severe positivity violations, A-TMLE demonstrates greater robustness in finite samples and provides narrower confidence intervals.

Beyond its improved performance in challenging settings like near positivity violations, A-TMLE has many exciting applications in data augmentation, such as integrating randomized controlled trial (RCT) data with observational real-world data (RWD) (van der Laan et al. [2024]). Combining RCT data with external RWD allows for more efficient and precise estimation of treatment effects, especially if the drug has only been approved in specific regions. In these settings, investigators can leverage data from patients outside of the trial. However, this hybrid design (i.e., data fusion or data integration) faces challenges to validity, including differences in the trial population versus the real-world population. In van der Laan et al. [2024], the authors proposed a method within the A-TMLE estimation framework that uses HAL to estimate a working model for the conditional effect of being enrolled in the RCT on the outcome of interest. This working model allows the borrowing of information from external RWD. Simulations have demonstrated that this method outperforms alternative data augmentation methods by achieving smaller mean squared error when estimating the ATE, especially when the bias is a simple function, such as constant bias. Unlike existing methods, the efficiency gain using this method is primarily driven by the complexity of the bias model rather than the magnitude of the bias. Asymptotically, it will always be at least as efficient as an efficient estimator that uses only the RCT data.

## 5 Code example

For practitioners who are interested in applying HAL to their research data to estimate parameters (including ATE, CATE, and dose-response curves) and construct valid statistical inference, we have provided a code example on GitHub (https://raw.githack.com/SeraphinaShi/HAL-in-R/main/Demo.html) that outlines how to (1) fit a HAL with cross-validation-selected penalty factor or an undersmoothed HAL, (2) interpret results from a HAL model, and (3) estimate target parameters with HAL as a working model and obtain delta method based confidence intervals.

## 6 Discussion

In this article, we discuss the theoretical advantages and various applications of Highly Adaptive LASSO in causal inference problems. HAL offers valid statistical inference by requiring only that the relevant functions of the statistical model are *càdlàg* with a bounded variation norm. HAL acts as a maximum likelihood estimator within this variation norm-constrained statistical model, where the variation norm can be selected via cross-validation. When interested in functional parameters, such as a conditional mean rather than the entire data density, HAL acts as an empirical risk minimizer over the set of *càdlàg* functions with bounded sectional variation norms.

HAL possesses several important theoretical properties: 1) a dimension-free rate of convergence in terms of loss-based dissimilarity; 2) asymptotically efficient plug-in estimators of pathwise differentiable target parameters when undersmoothing is applied; and 3) pointwise asymptotic normality as an estimator of the target function itself. The first property is crucial for establishing asymptotic linearity and efficiency of estimators such as TMLE. In particular, the analysis of these estimators depends on an exact remainder being negligible at a rate of *n*^−1*/*2^, which necessitates that the nuisance functions are estimated at a rate faster than *n*^−1*/*4^. Consequently, HAL is a perfect candidate machine learning algorithm that not only guarantees consistency but also approximates the true target function at the required rate as sample size increases. The second property allows HAL to function as a plug-in estimator independently of TMLE or other efficient influence curve-based debiasing techniques. This is particularly useful when the investigator is not familiar with deriving canonical gradients, as it enables the construction of asymptotically linear and efficient estimators without needing to derive them. Finally, the third property enables HAL to provide valid inferences for non-pathwise differentiable parameters, such as the causal dose-response curve, further expanding its applicability in causal inference.

HAL’s limitation lies in its high memory and computational requirements. Specifically, using LASSO software like the *glmnet* R package for optimization requires augmenting the design matrix, which can grow as large as *n ×* 2^*d*−1^, placing a significant burden on both memory and computation. To mitigate this issue, one can tune HAL’s hyperparameters by limiting the maximum number of interactions (e.g., restricting basis functions to 3-way interactions) or reducing the number of knot points (e.g., selecting a smaller number of knot points based on quantiles or using clustering algorithms like *k*-means to select centroids as knot points). Despite these strategies, developing more computationally efficient solutions for HAL remains an active area of research.

## Data Availability

All data used in and produced by the present study are available upon request.

## 7 Acknowledgments

## 7.1 Funding

This research was funded by the National Institute for Allergy and Infectious Diseases (1K01AI182501-01 to Zachary Butzin-Dozier) and a global development grant (OPP1165144) from the Bill and Melinda Gates Foundation to the University of California, Berkeley, CA, USA.

## Notes

### Competing Interest Statement

The authors have declared no competing interest.

